# Infection Inspection: Using the power of citizen science to help with image-based prediction of antibiotic resistance in *Escherichia coli*

**DOI:** 10.1101/2023.12.11.23299807

**Authors:** Alison Farrar, Conor Feehily, Piers Turner, Aleksander Zagajewski, Stelios Chatzimichail, Derrick Crook, Monique Andersson, Sarah Oakley, Lucinda Barrett, Hafez El Sayyed, Philip W. Fowler, Christoffer Nellåker, Achillefs N. Kapanidis, Nicole Stoesser

## Abstract

Antibiotic resistance is an urgent global health challenge, necessitating rapid diagnostic tools to combat its escalating threat. This study introduces innovative approaches for expedited bacterial antimicrobial resistance profiling, addressing the critical need for swift clinical responses. Between February and April 2023, we conducted the Infection Inspection project, a citizen science initiative in which the public could participate in advancing an antimicrobial susceptibility testing method based on single-cell images of cellular phenotypes in response to ciprofloxacin exposure. A total of 5,273 users participated, classifying 1,045,199 images. Notably, aggregated user accuracy in image classification reached 66.8%, lower than our deep learning model’s performance at 75.3%, but accuracy increased for both users and the model when ciprofloxacin treatment was greater than a strain’s own minimum inhibitory concentration. We used the users’ classifications to elucidate which visual features influence classification decisions, most importantly the degree of DNA compaction and heterogeneity. We paired our classification data with an image feature analysis which showed that most of the incorrect classifications were due to cellular features that varied from the expected response. This understanding informs ongoing efforts to enhance the robustness of our deep learning-based bacterial classifier and diagnostic methodology. Our successful engagement with the public through citizen science is another demonstration of the potential for collaborative efforts in scientific research, specifically increasing public awareness and advocacy on the pressing issue of antibiotic resistance, and empowering individuals to actively contribute to the development of novel diagnostics.

**Lay summary:** Antibiotic resistance is a big health problem worldwide. We need fast ways to find out if bacteria are resistant to antibiotics. In our study, we develop new methods to do this quickly. We ran an online project called Infection Inspection from February to April 2023, in which 5,273 people took part. Together, they classified more than a million pictures of bacterial cells, helping our project use these pictures to detect antibiotic resistance. The volunteers performed well, getting near 67% of the answers right. We also learned which pictures helped or confused them. This will help us make our computer program better. This project didn’t just help science; it also taught people about antibiotic resistance. Partnerships between the public and scientists can make a difference to developing technologies that protect our health.

## Introduction

Antibiotic resistance is an escalating global health concern, necessitating the development of new technologies such as rapid tests for antibiotic-resistant bacteria to mitigate its impact. Rapid identification of which bacterial species is causing an infection and its resistance profile has been shown to both optimize antibiotic use and enhance patient outcomes^1,2^. Currently, typical diagnostic tests rely on time-consuming bacterial culture growth, taking a minimum of 12 to 48 hours to produce results. Alternative rapid tests focus on identifying resistance-associated genes, but these may not always directly correlate with phenotypic resistance^3^. Antibiotic resistance poses a significant threat to individual and public health by potentially rendering common antibiotics ineffective in treating bacterial infections, but public awareness of the use of antibiotics and the impact of antibiotic resistance remains incomplete^4^.

Citizen science collaborations between volunteers and research teams can play an important role in educating the public about scientific concepts and have been instrumental in recognizing complex patterns within biological data, starting with research in ecology and extending to various biological fields, including protein folding, DNA sequence alignment, electron microscopy, and microbiology^5–7^. These projects enable individuals of diverse backgrounds and expertise levels to contribute to scientific data collection and analysis, empowering them to actively advance and acquire knowledge in various disciplines. Public involvement broadens the spectrum of available data, perspectives, and ideas, leading to more comprehensive and innovative research outcomes. Successful examples, such as the Great Backyard Bird Count and the Merlin Bird ID app^8,9^, demonstrate citizen science’s potential to enable large-scale data collection and analysis, raise awareness, inspire future scientists, and promote environmental and civic responsibility.

The public can be effectively engaged in citizen science projects using various strategies, including hosting events, utilizing social media platforms, partnering with educational institutions and community organizations, and offering training and educational resources, but most of these approaches engage only 10s-100s of individuals. Online platforms which simplify access for large-scale, global public engagement in targeted or diverse citizen science projects include Zooniverse^10,11^, SciStarter^12^, and Foldit^13^ amongst others^14^. For participants, Zooniverse offers a unique and engaging way to learn about science, participate in real research, and connect with like-minded individuals^15^.

A previous project hosted on Zooniverse, Bash the Bug^7^, successfully engaged citizen scientists to look at images of bacterial growth and identify their resistance to antimicrobial drugs. This demonstrated how citizen science can be used for antimicrobial resistance research and the development of novel diagnostic tools. We are developing a diagnostic method that relies on a microfluidic device for the direct capture and identification of bacteria and associated antibiotic resistance from clinical samples using microscopy. We recently developed a deep-learning model which can classify individual *E. coli* cells as ciprofloxacin-sensitive or resistant with 80% accuracy (which results in high-confidence classifications of populations of bacteria) based on morphological changes to the sub-cellular structure^16^. The continued development of these single-cell, imaging-based classification methods requires robustness to bacterial heterogeneity, and an understanding of why certain cells within a sample are misclassified is essential. We therefore developed a project on Zooniverse called Infection Inspection to leverage the power of citizen scientists towards optimising our novel method, and to engage the public in an antibiotic resistance-focused project. We first trained volunteers to recognize cellular phenotypes associated with ciprofloxacin-sensitive and ciprofloxacin-resistant *E. coli*, and then used their classifications to learn what features facilitate accurate classification, and which lead to ambiguity and misclassifications. Our aim was to use their classifications and misclassifications to make our machine learning-based bacterial classifier more robust to atypical phenotypes, whilst simultaneously educating the citizen scientists about antibiotic resistance.

## Methods

### Image dataset

The project dataset was made up of 49,074 individual images of ciprofloxacin-treated *E. coli* cells generated for previous work^16^. All bacteria had been chemically fixed and stained using 4′,6-diamidino-2-phenylindole (DAPI) as the nucleic acid stain and Nile Red as the membrane stain. The initial dataset was composed of 11,074 256×256 Red-Green-Blue (RGB) images of *E. coli* cells from 5 clinical strains (EC1, EC2, EC3, EC5, and EC6, reported previously^16^) treated at 10 mg/L for 30 minutes, with clinical strains defined as ciprofloxacin-resistant (minimum inhibitory concentration [MIC] >0.5mg/L) or ciprofloxacin sensitive (MIC ≤0.25 mg/L) using European Union Council on Antimicrobial Susceptibility Testing (EUCAST) breakpoints^17^ . A second dataset of 38,000 images included the same *E. coli* strains treated at 9 concentrations ranging from 16 mg/L to 0.001 mg/L ciprofloxacin for 30 minutes. All bacteria were imaged in an automated workflow as agarose-mounted samples in phosphate buffered saline (PBS) on a Nanoimager-S fluorescence microscope (Oxford Nanoimaging) using the multiple acquisition capability of the microscope with autofocusing on each field of view. The image segmentation for background removal was done with an optimised model of Mask-RCNN adapted from a standard implementation^18,19^.

### Development of Infection Inspection with the Zooniverse project builder workflow

Infection Inspection was designed as a citizen science project on the Zooniverse platform (https://www.zooniverse.org/) using the Project Builder (https://zooniverse.org/lab), a free-to-use web browser application enabling research teams to build and contribute projects to the site. During the building process, we developed an initial workflow, tutorial, and project field guide. Datasets were added to the project as .png images using the Subject Set upload tool within the Zooniverse Project Builder.

Infection Inspection was submitted for internal review in August 2022 and went to beta reviewers in September 2022. In response to beta feedback, we improved our project terminology, added explanations to the field guide and instructions for how to classify ambiguous or unusual cells. During the beta test, we noticed that user accuracy did not improve with the number of classifications done. To help users learn from their own misclassifications, we implemented user feedback for a set of 30 tutorial images. These images had a ground truth classification of “Sensitive,” “Resistant,” or “Image Processing Error” and users would receive feedback on their accuracy immediately after submitting a classification for one of these images. Tutorial images were shown to users with decreasing probability: 0.5 for the first 5-10 subjects, declining to 0.25 by 20 subjects, and 0.05 after 50 subjects. The retirement limit was set to 20, meaning that each image was considered complete once it was classified by 20 unique volunteers.

The tutorial and field guides were written in line with guidance on communicating with the public on antibiotic resistance from the Wellcome Trust^20^. We solicited and implemented feedback from non-experts, public engagement experts, and a secondary school biology teacher on the language used in the project before submitting for beta testing.

### Accuracy and participation analysis

On completion of the project on 10^th^ May 2023, the project data file which included all classifications was downloaded in .csv format from the Zooniverse site. Only classifications performed from go-live (7^th^ Feb 2023 17:40 UTC) to full dataset completion (10 May 2023 21:40 UTC) were included in the analysis. Image identifiers were matched back to the original strain and metadata including known MIC, treatment concentration, clinical antibiotic susceptibility phenotype, and predicted classifications were assigned to each data point. The predicted classification was defined as the expected response of the strain for the antibiotic. For instance, if the MIC for a particular strain was 0.03 mg/L and the treatment concentration was 10 mg/L, that strain was categorized as susceptible. Conversely, if the MIC was 72 mg/L and the treatment concentration was 10 mg/L, the strain was labelled as resistant. All usernames were anonymised to ‘User_1, _2, etc’. For the accuracy determination, any classifications of images that were part of the training/feedback dataset or classifications of “Image processing error” were removed. Summary statistics were performed in R (v 4.2.3) using the R package vegan (v 2.6-4) and plotted with ggplot2 (v 3.4.4). Accuracy was graded as whether the user’s classification matched the image’s predicted classification as defined above.

We used the Gini coefficient to characterise the extent of inequality in the distribution of classifications by volunteers. The Gini coefficient derives from a metric for income inequality^21^ and has been applied to measure inequality in volunteer contributions previously^15^. We calculated the Gini coefficient with the following formula:

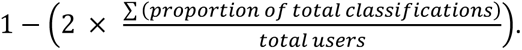

### Image feature analysis

CellProfiler^22^ was used to extract image features from the dataset. The RGB .tif images were split into grayscale single-color images using the ColorToGray module. Then, the IdentifyPrimaryObjects module was used with default settings to identify the Membrane object from the red channel. For the Nucleoid object, two-class Otsu thresholding was used with default settings because it segmented diffuse nucleoid regions more accurately. Intensity measurements for each object were measured using the MeasureObjectIntensity module. Size and shape measurements were extracted using the MeasureObjectSizeShape module. All measurement data were exported using CellProfiler to an SQLite database^23^ and selected measurements were converted to .csv files with DB Browser for SQLite (version 3.12.2).

Further image feature analysis was completed in Python and R scripts, available at https://github.com/KapanidisLab. Images were excluded from analysis if more than half of their classifications were “Image Processing Error.” An accuracy threshold of 0.5 was chosen to compare images that were most frequently classified correctly or incorrectly. For example, if a cell’s predicted classification was Sensitive, based on its MIC and the treatment concentration, and it was classified as Sensitive by more than 50% of users, it would be labelled as Correct Sensitive. A cell from the same strain and treatment condition that was classified as Resistant by more than 50% of users would be labelled as Incorrect Sensitive. This yielded four sets of images whose features could be compared: Correct Sensitive, Correct Resistant, Incorrect Sensitive, and Incorrect Resistant. Images were called Most Correct if they were classified correctly with a ratio greater than 0.94, corresponding to roughly 19 correct classifications of 20.

For feature comparisons between groups of cells displaying ciprofloxacin-resistant or susceptible phenotypes, we performed two-sided *t*-tests with Bonferroni corrections for multiple comparisons using the ggpubr (version 0.6.0) and Rstatix (version 0.7.2) packages. We compared the distributions of the values associated with cellular phenotypic features to a normally distributed Random Noise feature generated by numpy.random.normal^24^. A principal component analysis with 2 principal components was performed using the 7 measured image features and the Scikit-learn PCA function^25^. Before analysis, all feature measurements were normalised with Standard Scaler from Scikit-learn^25^. For the dataset with multiple ciprofloxacin concentrations, the principal component analysis with 2 principal components was performed in R with the prcomp function from the stats library (version 4.1.3) and plotted with ggplot2 (version 3.4.3).

Independently, we extracted the feature importance values from a Random Forest classifier with 100 trees and a minimum of 3 samples per leaf that had been trained using Scikit-learn^25^ on images that were randomly allocated to a 75-25 train-test split and then scaled with Standard Scaler. The Random Forest classifier was evaluated by cross-validation by the Mean Absolute Error and was then applied to the test dataset to make predictions.

For each image, we calculated SHAP (SHapley Additive exPlanation) values for each feature using Kernel SHAP, a model agnostic implementation for Python^26^. This method, which derives from game theory approaches, measures an importance value for each feature for each image classification, and has been shown to correspond well to intuitive human feature impact estimates^26^.

## Results

### The Infection Inspection project engaged a large cohort of users

Infection Inspection was launched on the Zooniverse platform on the 7^th^ of February 2023 (Fig. 1a) and was promoted via multiple platforms, including webpages, print magazines, in-person outreach events, and emails for Zooniverse users. An initial dataset comprising of 30 training and 5,000 test images was available to users and completed within just 18 hours. A second dataset of 6,074 images was subsequently uploaded and completed in 72 hours, whilst a final dataset of 38,000 images was uploaded and completed in 35 days (840 hours). A total of 5,273 unique users performed at least one image classification and overall 1,045,199 classifications were made between the project launch date and May 10^th^, 2023. After removing classifications of the training dataset, a total of 4,927 users remained, covering 1,003,588 classifications (Fig. 1b). The median number of classifications performed by users was 38, however the variation in number of classifications per user was large, with 56 users performing >2,000 individual classifications. The maximum classifications undertaken by any given user was 46,289.

**Figure 1.**
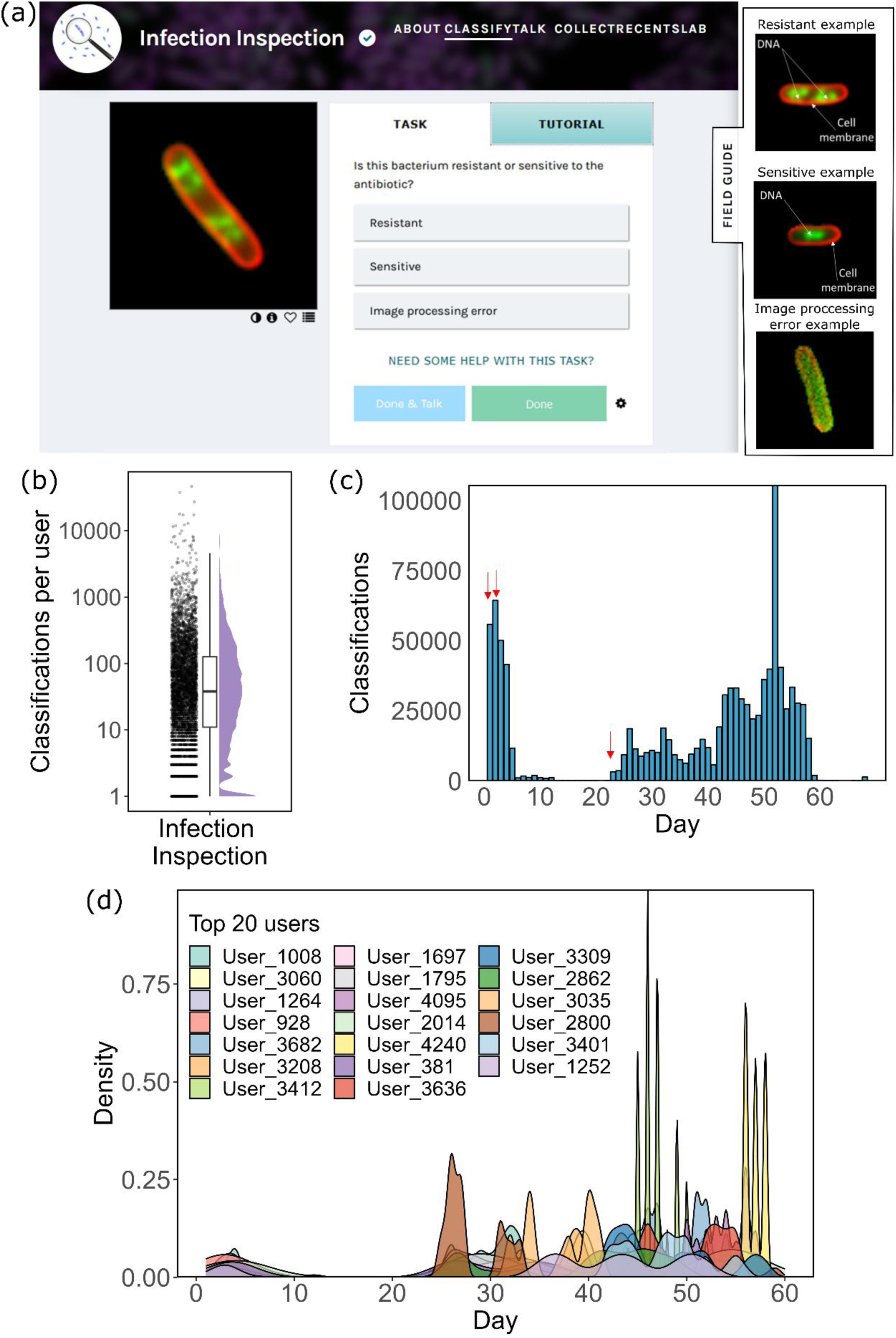
Engagement of users with Infection Inspection. (a) Task page for users of Infection Inspection. Users were presented with an image of a bacterial cell and were asked to select from one of three options to classify the image. Accessing the field guide provided examples of each cell type as exampled in the subset panel (b) Distribution of the number of classifications by user (n=5273 users, n=1,045,199 classifications). Each dot represents an individual user that performed a classification on at least 1 non-training set image. The box represents the middle 50% (IQR) of the users and mid-line indicates the median number of classifications. (c) The distribution of classifications performed on each day of the project. Red arrows indicate the time of each data batch upload. (d) Density mapping of activity for each of the top 20 users (by number of classifications) over the course of the project highlighting the differences in patterns of contribution. Day 1 on the x-axis represents the first day that the user engaged with the project.

Engagement with the project correlated with the upload of new data, with spikes in classifications occurring within 1-2 days after upload (Fig. 1c). Amongst the 20 most engaged users, return to the project was common, with these users returning to the project on several occasions throughout (Fig. 1d). The Gini coefficient for our user participation was 0.81, which means that the most prolific volunteers contributed a large proportion of our project’s classifications, or our project attracted many casual users, or both. The Gini coefficient for Infection Inspection is close to the mean Gini coefficients of the most popular ecology (0.80), astronomy (0.82), and transcription (0.81) projects on Zooniverse and higher than the average Biomed project score of 0.67 based on a previous analysis^15^.

### Volunteers classified *E. coli* cellular phenotypes with accuracy comparable to deep learning

We assessed the accuracy of user classifications in distinguishing bacteria as either ciprofloxacin-resistant or susceptible, based on the ciprofloxacin treatment concentration relative to the Minimum Inhibitory Concentration (MIC) for each strain. When we aggregated the data from all three dataset uploads, users achieved an accuracy of 66.4% in classifying susceptible cells (Fig. 2a). The accuracy for classifying resistant cells was similar, standing at 67.3% (Fig.2a). We also employed the same images to test a deep-learning model^16^. Compared to the volunteers, the model was less accurate in classifying resistant cells (62.5%; Fig. 2a), but more accurate in classifying susceptible cells (88.2%).

**Figure 2.**
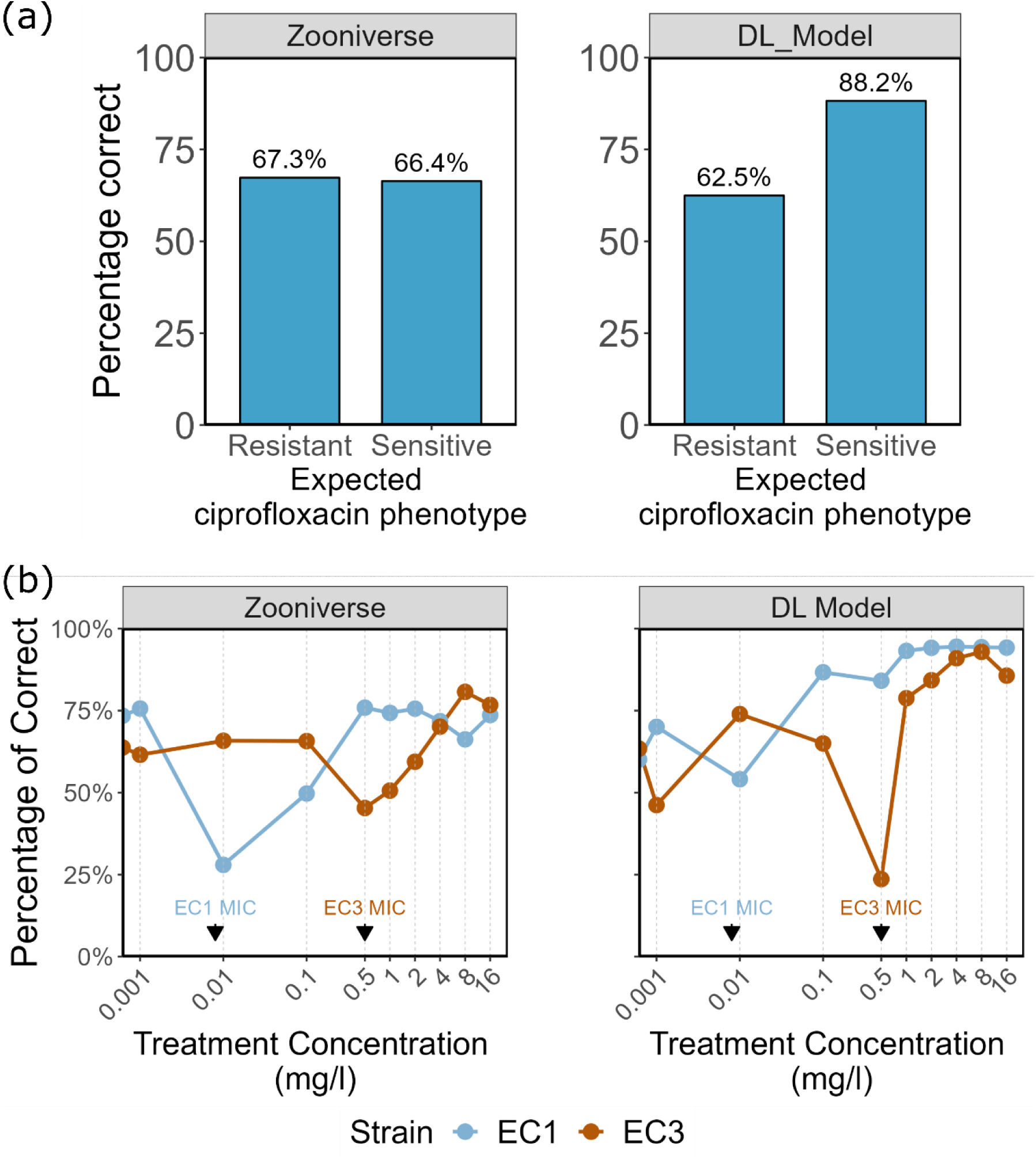
Classifications of images by Infection Inspection users. (a) An aggregated plot of how accurate users or model were at categorising an image into either the resistant or sensitive category based on the expected phenotype for the given sample. DL model denotes the deep learning model applied. (b) The line plots visualise the percentage of images that were correctly categorised as either resistant or susceptible as expected based on the treatment concentration, by the users or model. Each subplot shows the data for a different *E. coli* strain with the treatment concentration on the x-axis. The known, predetermined MIC for each strain is indicated on the plot using arrows.

Given that the number of classifications performed by users varied, we examined whether there was a correlation between accuracy and the total number of images classified. Despite having ten users who classified >10,000 images, we observed no significant relationship between accuracy and the total number of images classified (Fig. S1a). Additionally, the duration of user activity on the project did not influence classification accuracy (Fig. S1b).

### Classification accuracy depended on the antibiotic concentration used for treatment

In the third dataset uploaded, we introduced cells treated at varying concentrations of ciprofloxacin; some of the concentrations used were below the MIC, and thus we expected to see no significant phenotypic changes; on the other hand, some of the concentrations were above the MIC, and should produce phenotypic changes. These treatments allowed us to investigate whether both users and our deep learning model could detect changes in cell structure based on a graduated treatment concentration. For one of the strains (EC1), users correctly classified the cellular changes close to 75% of the time for all treatment concentrations except the one closest to the known MIC of the strain (Fig. 2b). At this specific treatment concentration (0.01 mg/L), accuracy in identifying the response dropped to nearly 25%. A similar trend was observed in the model’s predictions. While accuracy was highest at treatment concentrations of 0.1 mg/L and above, the greatest confusion was encountered when the treatment concentration approached the MIC of the strain (Fig. 2b). This pattern of increased confusion was also observed for the second strain (EC3), which had a different MIC (0.5 mg/L).

### Differences in DNA morphology leads to the most confusion in correctly classifying images

Some images were more frequently misclassified than others. In the first uploaded dataset of 3,015 ciprofloxacin-sensitive and 3,212 ciprofloxacin-resistant cells treated at 10 mg/L, the classification accuracy histograms are left-skewed, with many images almost always classified correctly, and others almost never (Fig. 3). This suggested that, while many cells displayed the expected cellular phenotype when exposed to ciprofloxacin, there were sub-populations with atypical features.

**Figure 3.**
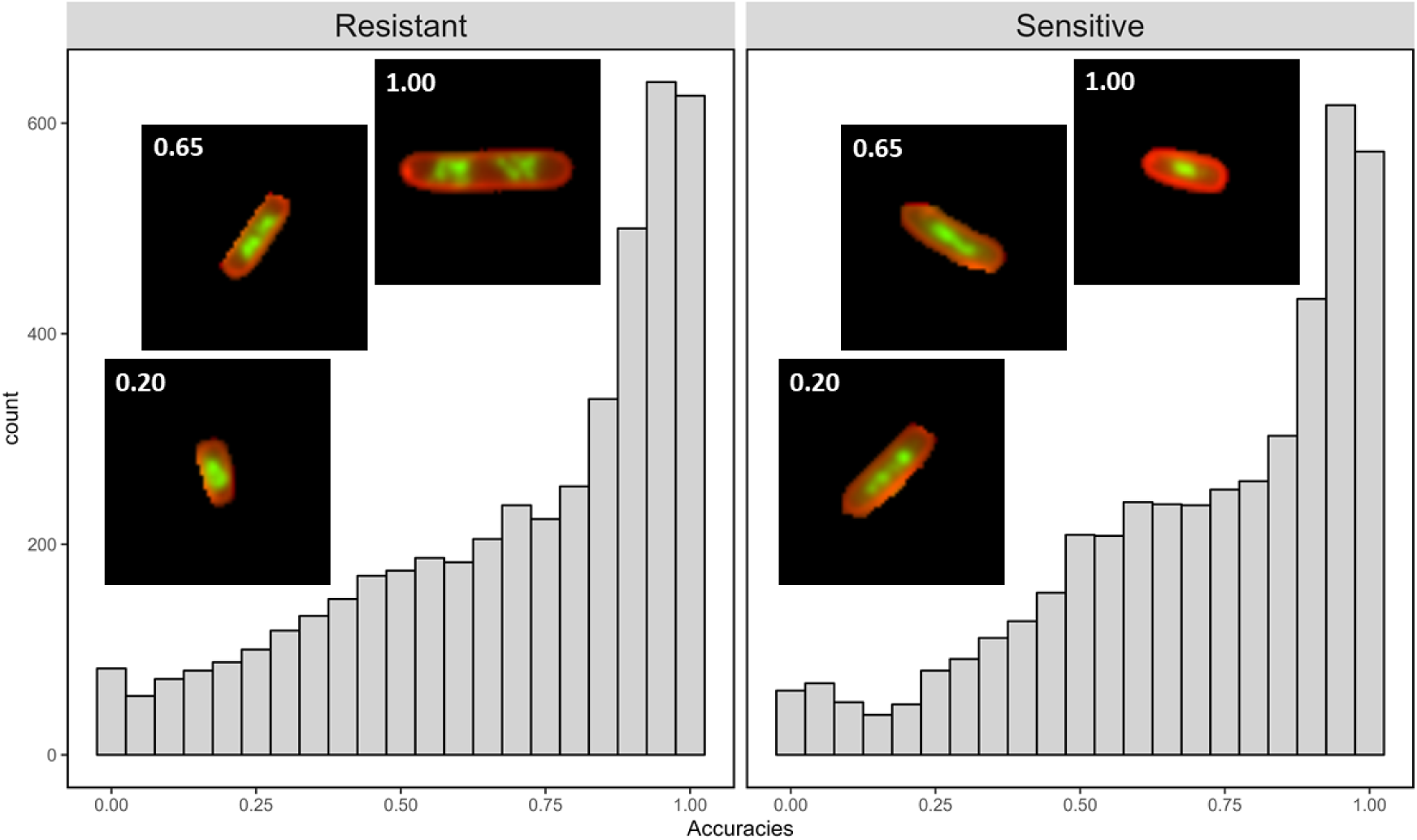
User accuracy varies by image for both resistant and sensitive cells. Histograms of the user accuracy on images of *E. coli* treated at 10 mg/L for resistant and sensitive cells. Representative images of resistant and sensitive cells with low classification accuracy (0.20), intermediate classification accuracy (0.65) and high accuracy (≥0.95) are shown. Both resistant and sensitive cells show a left skew, with many cells being classified correctly nearly always and some almost never. However, both populations also have many ambiguous images that were classified correctly by around half of the users.

For this specific analysis, images were assigned to the “Incorrect” class if they were classified with less than 50% accuracy, and otherwise to the “Correct” class. Cells were excluded from the image feature analysis if they labelled as an “Image Processing Error” by more than half of the users who classified the image; this removed 230 images.

To explore why some cells were more frequently misclassified than others, human-interpretable image features were measured with CellProfiler^27^. Seven features were chosen for their potential biological relevance to the ciprofloxacin response (Fig. S2). To characterize the compaction, heterogeneity, and quantity of DNA, we measured the number of DNA regions per cell, the mean and standard deviation of the integrated intensity of the DNA regions, the mean standard deviation of the DNA intensity, and the area fraction occupied by the nucleoid regions. The cell shape was described by the form factor of the membrane and the major axis length was used to measure the cell size.

The image features of susceptible and resistant cells that were most often classified correctly were all significantly different (Corrected *t-*test p<0.001) (Fig. S2). However, when comparing the features of cells that were most frequently classified incorrectly, there was no significant difference in the mean integrated intensity of the DNA regions (p=1), the mean standard deviation of intensity of the DNA regions (p=0.34), and Nucleoid Area Fraction (p=1) between sensitive and resistant bacteria (Fig. 4), consistent with the images of these cells having features that are too similar to distinguish.

**Figure 4.**
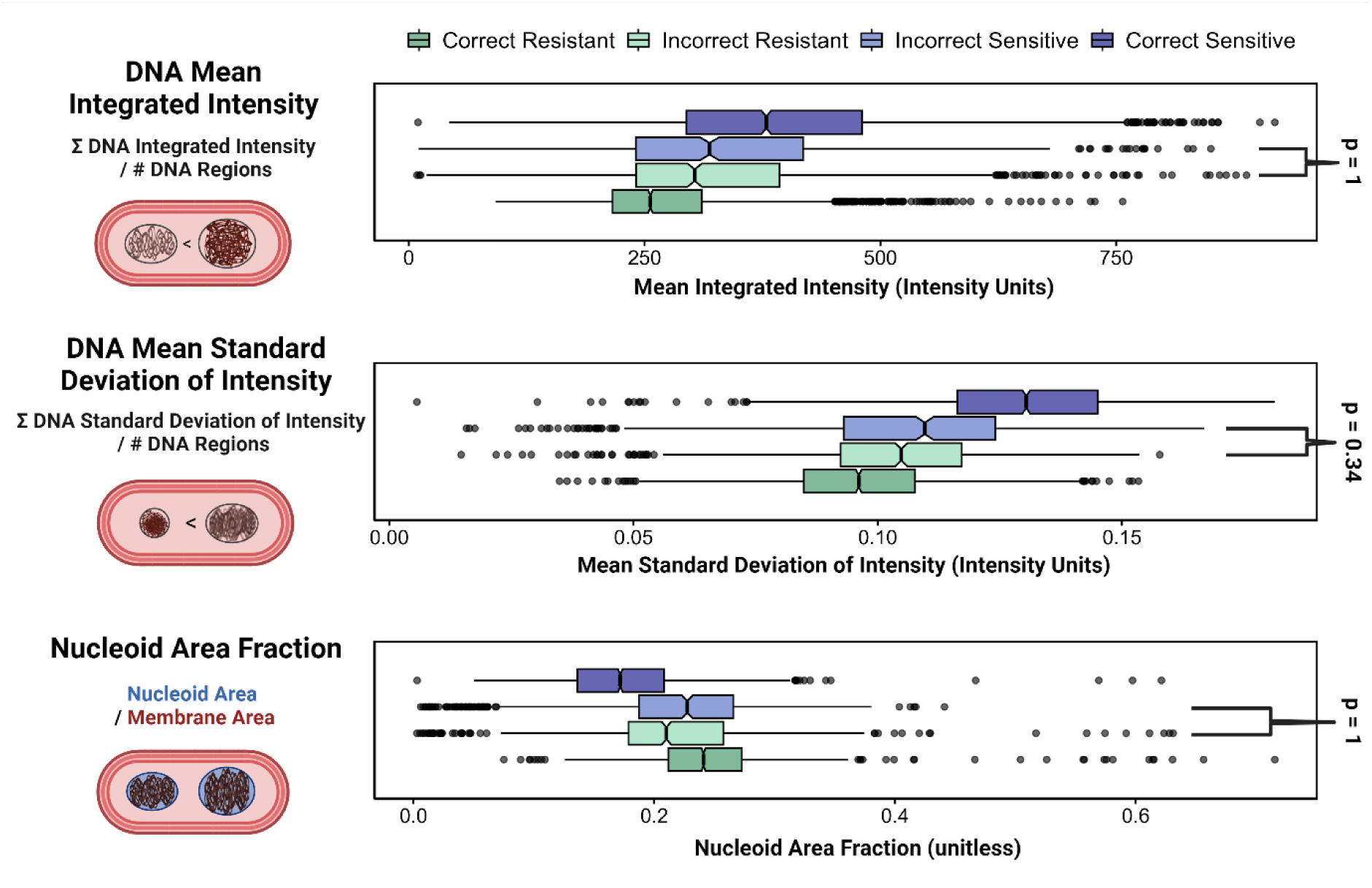
Cells that were classified incorrectly have more similar features. For three image features related to DNA heterogeneity and compaction, illustrations and definitions of which are shown above, the Incorrect Resistant and Incorrect Sensitive feature distributions are not significantly different, as shown in the box plots. A cell is called “Incorrect” if more than 50% of user classifications disagreed with the cell’s predicted classification, based on its MIC and the antibiotic treatment concentration. Notches are drawn showing the median value for each feature, and outliers are shown as spheres. The Bonferroni-corrected p-values were calculated for each pairwise comparison, and the features that were not significantly different are shown with brackets; all the other pairwise comparisons were significantly different with p < 0.0001.

### Images classified correctly and incorrectly cluster separately with distinct feature properties

To understand the cellular phenotypes represented by our image features, a principal component analysis was performed. The principal component analysis allowed us to visualise the phenotypic variance in our image dataset by projecting the feature measurements of each cell into a 2-dimensional space such that images with more similar features would cluster together. In addition, the loading vectors of each feature revealed the magnitude of its contribution to the ciprofloxacin-sensitive and ciprofloxacin-resistant phenotypes.

The variation in the first principal component was primarily driven by the number of DNA regions, the standard deviation of the integrated intensity of the DNA regions, and the cell major axis length; the second principal component was driven by the nucleoid area fraction and the mean integrated intensity of the nucleoid (Fig. 5). Images of susceptible or resistant bacteria that were in the correct class clustered separately, with some overlap, while images in the incorrect class clustered in the centre, with greater variation in the second principal component. This suggested that images that are frequently classified incorrectly have intermediate phenotypes, with DNA regions and cell lengths that were not clearly demonstrating signs of ciprofloxacin-resistance or sensitivity.

**Figure 5.**
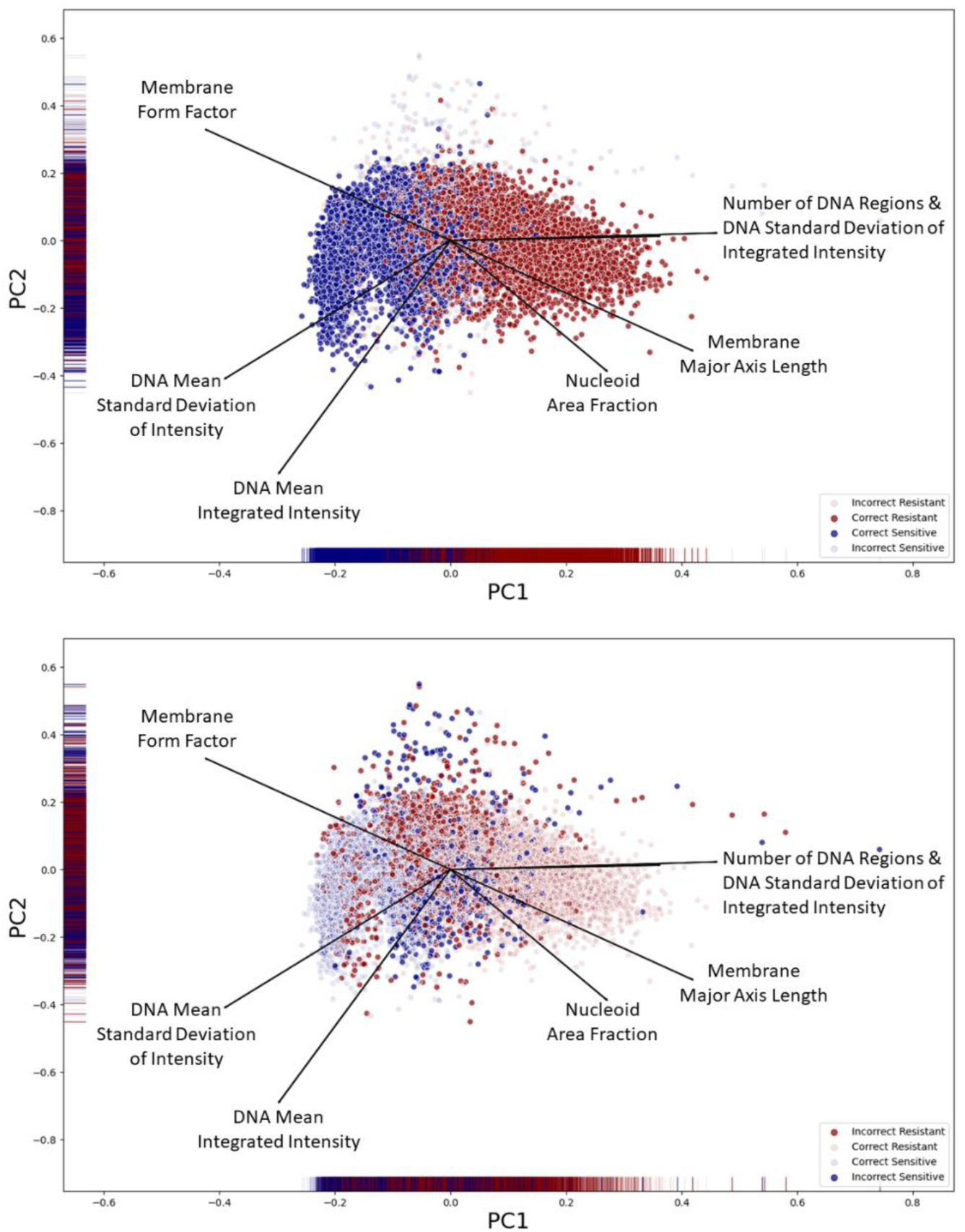
Incorrectly classified images have intermediate phenotypes. **Top:** In a 2-dimensional Principal Component Analysis, images of Resistant and Sensitive cells that were classified correctly more than 50% of the time (Correct Sensitive and Correct Resistant) cluster together, with some overlap. **Bottom**: Images that were classified incorrectly more than 50% of the time cluster near the centre of the principal component plot, with greater variance in the second principal component than correctly classified images.

For images where there was greater than 94% accuracy in classification (“Most Correct”), there was a distinct clustering observed with less overlap to the remaining correct images (Fig.S3). This highlighted that images are more likely to be consistently classified correctly when they exhibit features that distinguish them well from the opposite class.

### Ciprofloxacin sensitive and resistant cells portray different features of importance with respect to correct classification

We investigated which features might be most influential for classification (either by volunteers or a machine learning model) by computing SHAP contributions (SHapley Additive exPlanations)^26^. For the SHAP analysis, a Random Forest classifier was trained to classify images from our dataset as susceptible or resistant using our image feature measurements. An additional feature of normally distributed random numbers was added to determine which features held significance greater than random noise. This model achieved a Mean Absolute Error of 0.15 (accuracy = 85%) on a holdout dataset. The trained Random Forest model was then used to compute SHAP feature contribution scores for each image in the test holdout dataset. The average importance of a feature can be measured by the mean absolute value of the SHAP contribution for all images in the dataset.

Using this approach, and when looking at the entire dataset of susceptible and resistant images, the most important features were the DNA mean standard deviation of intensity (median SHAP=0.109, p<0.0001), number of DNA regions (median SHAP=0.085, p<0.0001), and nucleoid area fraction (median SHAP=0.081, p<0.0001) (Fig.6). All of the measured features contributed more to the classification task than the normally distributed random noise (p<0.0001).

**Figure 6.**
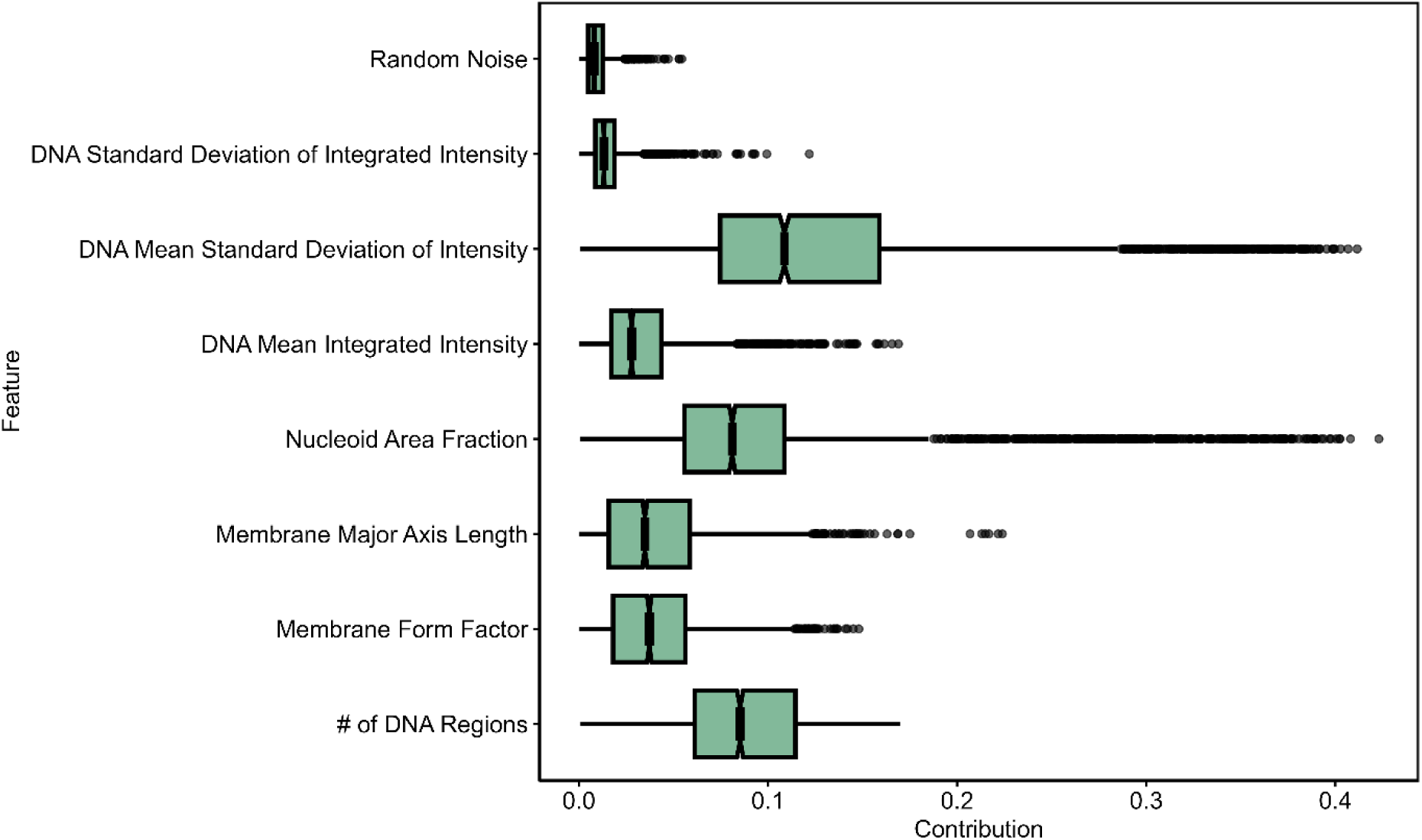
Feature significance for all images measured by SHAP values. The absolute value of the SHAP contribution for each feature is shown on a box plot, with *p*-values for each pairwise comparison with a Random Noise feature (*t*-test with Bonferroni correction for multiple comparisons). In order of their median SHAP contribution, the features are DNA Mean Standard Deviation of Intensity (0.109), Number of DNA Regions (0.085), Nucleoid Area Fraction (0.081), Membrane Form Factor (0.037), Membrane Major Axis Length (0.035), DNA Mean Integrated Intensity (0.028), DNA Standard Deviation of Integrated Intensity (0.013), and Random Noise (0.008). All the features are significantly different from Random Noise (p<0.0001), and all features are significantly different from each other (p<0.0001), except Membrane Major Axis Length and Membrane Form Factor (no significance).

### Different phenotypes develop in resistant bacteria treated with high concentrations of ciprofloxacin

In addition to the stark phenotypic differences between ciprofloxacin-treated susceptible and resistant bacteria treated at the same antibiotic concentration, our titration dataset revealed that an *E. coli* strain (EC3) with moderate resistance (MIC 0.5 mg/l) showed different features when treated at 8-, 16-, and 32-times the MIC (4, 8, and 16 mg/L, respectively) for 30 minutes compared to 2- and 4-times MIC (1 and 2 mg/l, respectively) (Fig. S4). This matched the trend in classification accuracy for EC3 at these concentrations (Fig. 2b).

## Discussion

The Infection Inspection project showed that misclassifications of ciprofloxacin-sensitive and ciprofloxacin-resistant *E. coli* are associated with diversity in the appearance of the bacterial DNA after antibiotic treatment. Ciprofloxacin is a fluoroquinolone antibiotic that inhibits the enzymes involved in bacterial DNA replication and repair^28^. In susceptible bacteria this can result in the compaction of the DNA and the inability to separate to dividing cells. Whilst our previously reported computer model could achieve a classification accuracy as high as 80%^16^ there remains a degree of confusion with respect to certain images, especially near the minimum inhibitory concentration of the strain (Fig. 2b). By image feature analysis, we found that images most likely to be classified incorrectly did not show the phenotypic features of correctly classified ciprofloxacin-susceptible or resistant cells, indicating that these bacteria develop ambiguous or intermediate phenotypes.

We used a feature analysis and computed SHAP contribution scores to determine that DNA mean standard deviation of intensity, the nucleoid area fraction, and the number of DNA regions were the most important features when deciding how to classify an image. This means that the degree of DNA compaction and heterogeneity, and the space it occupies within the cell, are the key features that can be used to determine whether an *E. coli* bacterium is responding to ciprofloxacin treatment.

The successful participation of the public with Infection Inspection and the speed at which users classified the images highlights the interest in and value of the public in tackling the problem of antimicrobial resistance. It is clear that citizen science platforms like the Zooniverse provide a valuable resource for recruiting large groups of the public to engage with research ^5,6^.

Our project demonstrates the utility of citizen science volunteers in interpreting large biomedical datasets. Biomedical projects are a minority on the Zooniverse platform. A 2019 study showed only 3 biomedical projects of 63 projects surveyed (5%)^15^ were included on the platform; as of November 13, 2023, this fraction remained low, with only 5/100 (5%) of active projects in a biomedical discipline. The Gini coefficient is a measure of inequality that has been used to assess the degree to which many casual volunteers and some super-users contribute to the shared work of Zooniverse projects. On average, biomedical projects were found to have a notably lower average Gini coefficient than astronomy projects, which could be the result of fewer return volunteers, or because biomedical projects more successfully attract many casual contributors^15^. Infection Inspection attracted 3,137 volunteers while it was active, with a Gini coefficient of 0.81, higher than the average biomedical project studied. We speculate that our single-step, fast workflow encouraged more classifications than the average biomedical research project.

Despite its successes, the Infection Inspection project had limitations. It relied on voluntary contributions from citizen scientists, which introduced variability in data quality and quantity. We had no information on the users participating in the study, and no quantitative feedback on the impact of our tutorials on informing the public about AMR. The study focused on a single antibiotic and cells obtained from a small number of bacterial strains, limiting the generalizability of findings to other antibiotics and pathogens. Since the workflow was limited to a single classification step, information about the volunteer’s decision-making process was lost.

Looking ahead, citizen scientists can continue to play a pivotal role in our research and in addressing global health challenges related to antibiotic resistance. Future engagements could involve exploring dynamic responses of bacterial cells to antibiotics, expanding the scope to cover a broader range of antibiotics and conditions, improving training materials and guidelines, raising public awareness, and integrating an assessment of the impact of these tools on user education about the scientific topics being studied. On the project discussion board, some volunteers started discussions about images that appeared to be cells in the process of cell division or images that looked unusual. While the project was not designed to classify images in such detail, it is encouraging to realise that that users could be asked to consider stages of cell growth in a future task. Our project, and other researchers working with citizen scientists, can take advantage of this scientific intuition in understanding their datasets.

In conclusion, the Infection Inspection project exemplifies the potential of citizen science platforms to engage the public in scientific research, enhance the analysis of large datasets, and contribute to our understanding of complex issues like antibiotic resistance. The collaboration between citizen scientists and researchers not only advances scientific methodologies but also fosters a sense of shared responsibility in addressing global health challenges. Despite its limitations, this project has opened doors to further exploration and collaboration, highlighting the promising role of citizen science in the future of biomedical research and public health.

## Data Availability

The raw data images used to build this project are available from the Oxford University Research Archive. The individual segmented single cell images and classification metadata are available at Zenodo.

https://ora.ox.ac.uk/objects/uuid:12153432-e8b3-4398-a395-abfb980bd84e

https://zenodo.org/doi/10.5281/zenodo.10301352

## Acknowledgements

The Zooniverse Volunteer Community: This publication was made possible by the contributions of volunteers in the Infection Inspection project. We thank them all for their dedication and engagement with the project. We would also like to thank the Zooniverse platform leaders, Helen Spiers, Mary Westwood, and Cliff Johnson. This work was supported by the Oxford Martin School (by the establishment of the Oxford Martin School Programme on Antimicrobial Resistance Testing; to A.N.K., N.S., C.N., D.C. and M.A.), by Wellcome Trust grant 110164/Z/15/Z (to A.N.K.), by the Clarendon Fund Scholarships (to A.F.), and by UK Biotechnology and Biological Sciences Research Council grants BB/N018656/1 and BB/S008896/1 (to A.N.K.). The research was additionally supported by the National Institute for Health Research (NIHR) Health Protection Research Unit in Healthcare Associated Infections and Antimicrobial Resistance (NIHR200915) at the University of Oxford in partnership with United Kingdom Health Security Agency (UKHSA) and by the NIHR Oxford Biomedical Research Centre. N.S. is an NIHR Oxford BRC Senior Research Fellow.

## Data availability

The raw data images used to build this project are available from the Oxford University Research Archive: https://ora.ox.ac.uk/objects/uuid:12153432-e8b3-4398-a395-abfb980bd84e. The individual segmented single cell images and classification metadata are available at: https://zenodo.org/doi/10.5281/zenodo.10301352.

---

**Figure S1.**
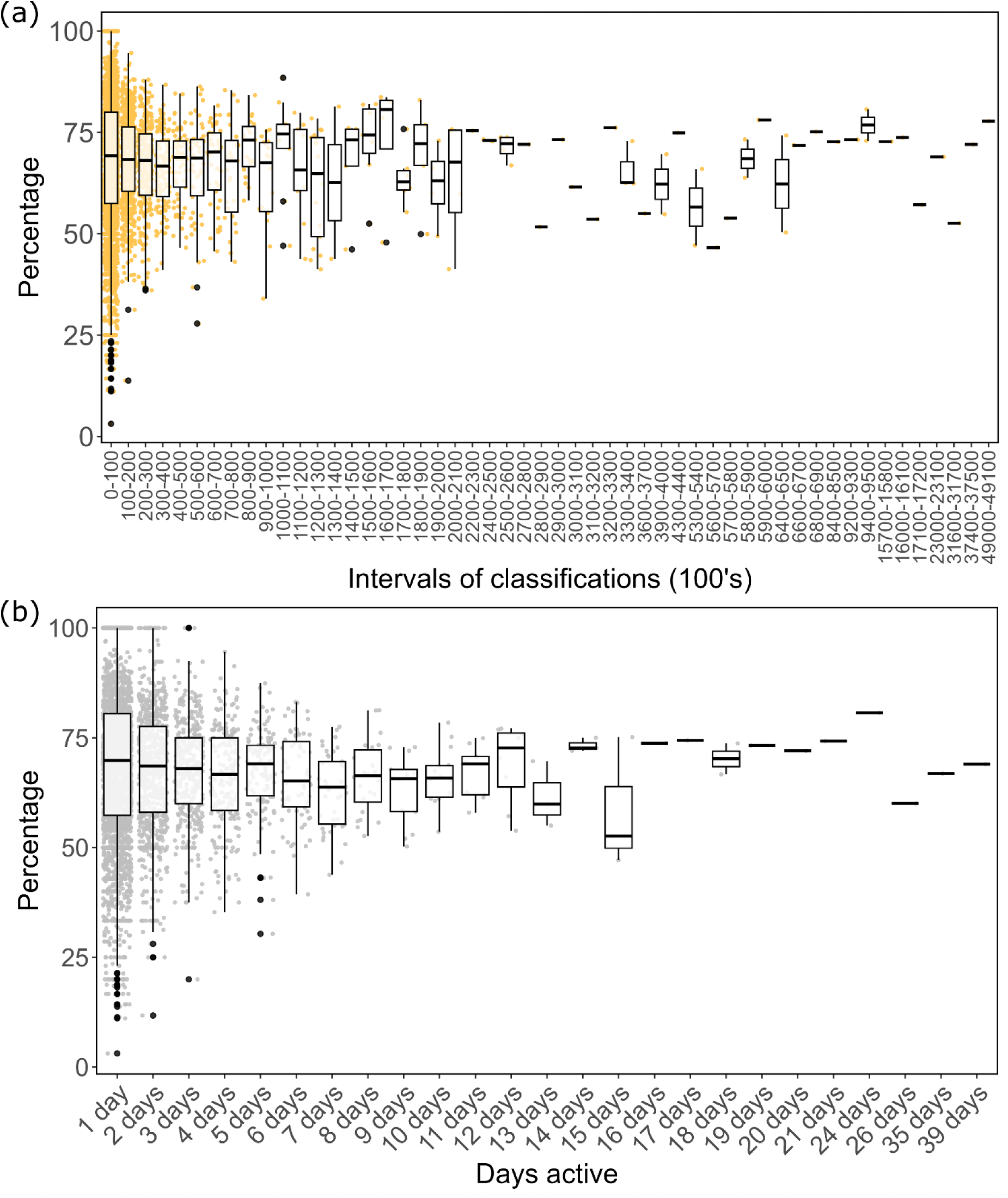
Accuracy of users based on engagement. Each boxplot represents the median image classification accuracy for users based on: (a) the total number of images they classified, or (b) total numbers of days they accessed the project. Boxplots highlight the middle 50% of data (IQR) with the median image classification accuracy for users shown in the central line.

**Figure S2.**
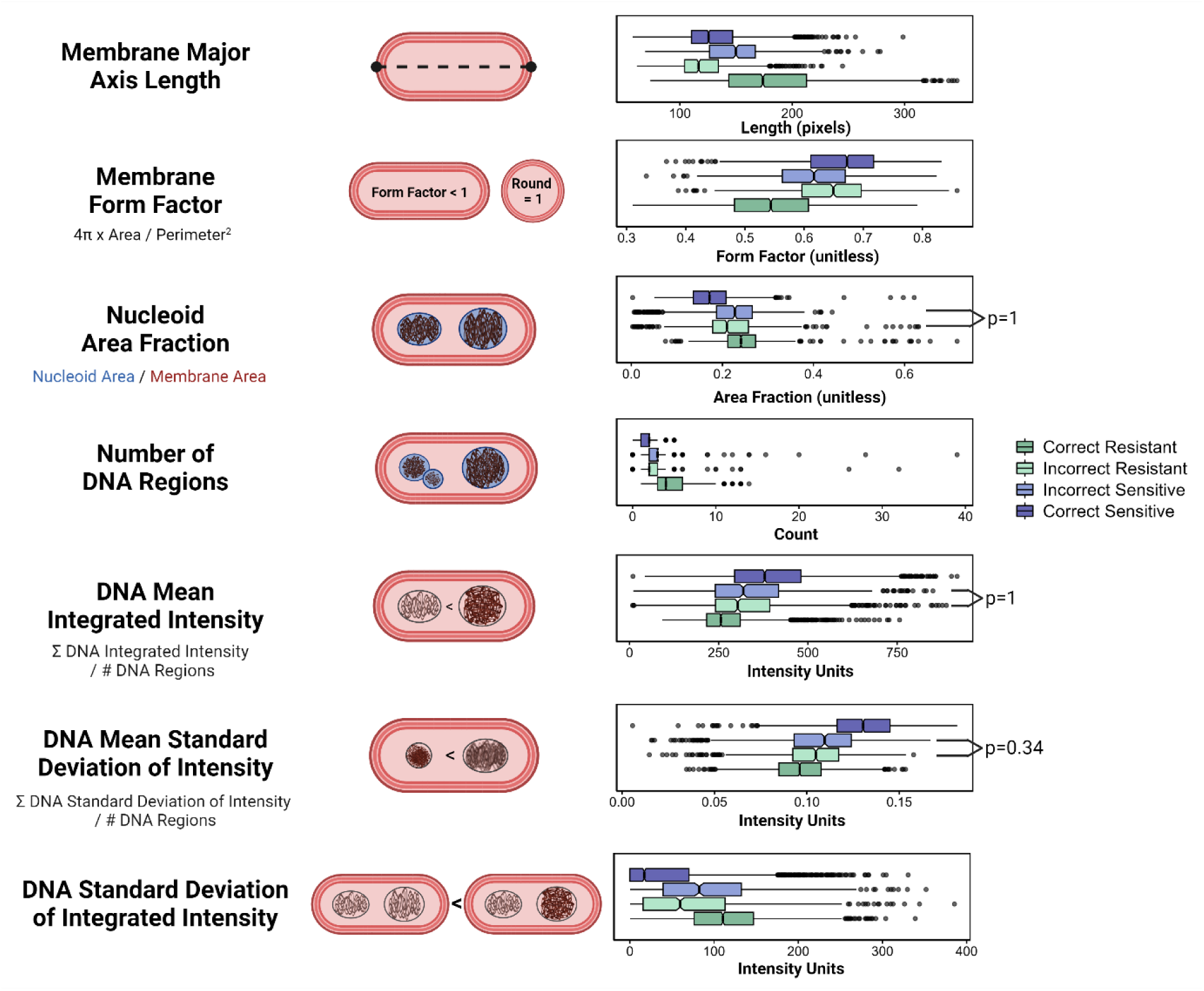
The definition and distributions of the seven measurements used for image feature analysis. Seven measurements were chosen for their potential to reflect responses to ciprofloxacin treatment. The features, with illustrative diagrams, are shown on the left; box plots of the feature distributions for all cells are shown on the right, coloured by whether the cell was Sensitive or Resistant and whether they were most often classified Correctly or Incorrectly. Notches indicate the median value and outliers are plotted as black dots. The Membrane Major Axis Length and the Membrane Form Factor measure cell size and cell shape, respectively. The Form Factor of a perfectly round object is equal to 1, so most bacilli will have form factors <1. The Nucleoid Area Fraction is a measurement of DNA compaction, DNA size, and cell size. The Number of DNA Regions detected by CellProfiler also reflects DNA compaction and cell cycle stage. Other measurements of the nucleoid, such as the Mean Integrated Intensity, Mean Standard Deviation of Intensity, and Standard Deviation of the Integrated Intensity of DNA regions, measure the changes in DNA heterogeneity and compaction as *E. coli* respond to ciprofloxacin. These measurements also capture the variations in nucleoid morphology that can be seen within the same cell. For each feature, pairwise t-tests were performed for the Correct Resistant, Incorrect Resistant, Incorrect Sensitive, and Correct Resistant distributions. The p-value with Bonferroni correction for multiple comparisons is <0.0001 except where labelled.

**Figure S3.**
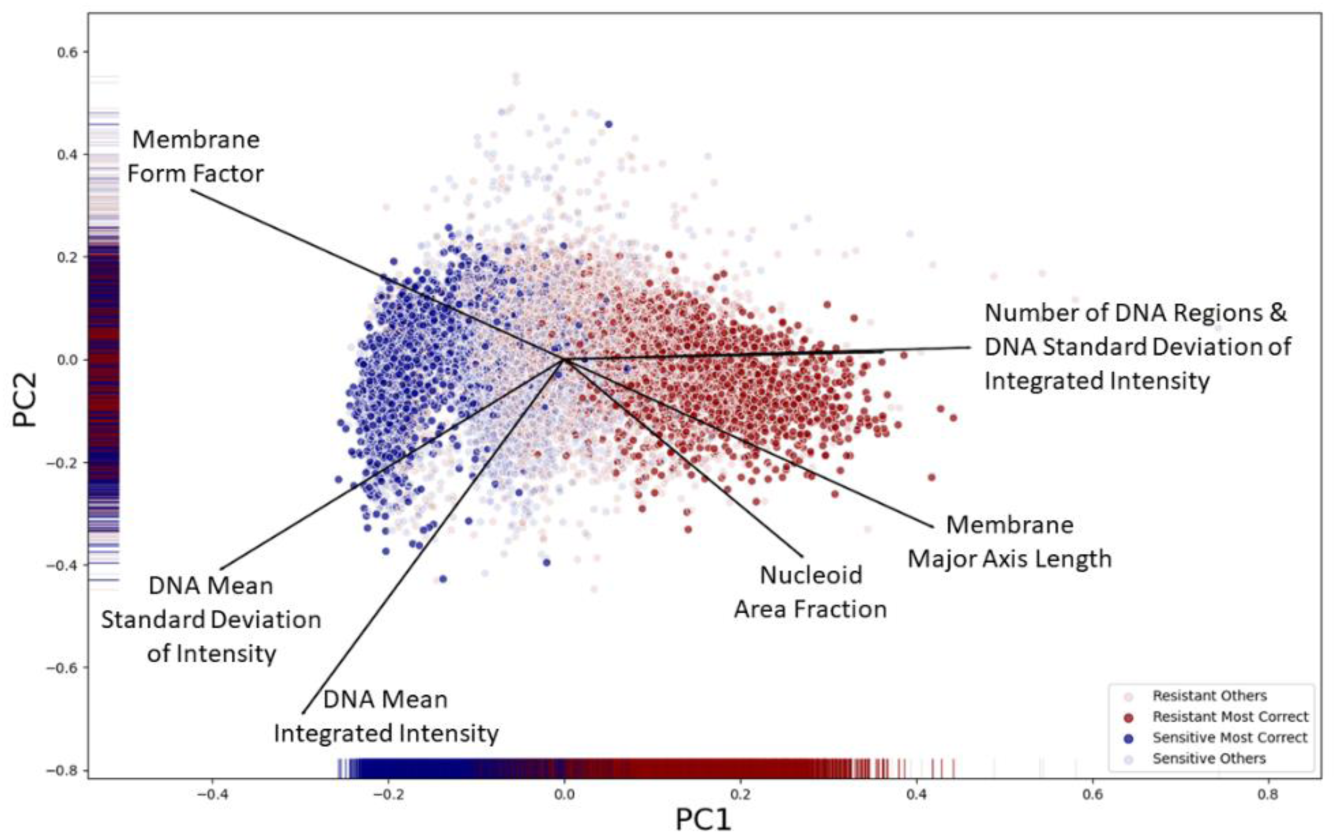
Features distinguishing the most correctly classified cells. Images were considered part of the Most Correct dataset if they were classified with greater than 94% accuracy (e.g. more than 19 times out of 20). The most influential features for the first Principal Component are the Number of Nucleoids and the Nucleoid Standard Deviation of Integrated Intensity, a measure of the variation in nucleoid region brightness within the cell. The Most Correct Resistant cells and Most Correct Sensitive cells form distinct clusters, indicating that there are certain populations of cells that exhibit characteristic Resistant and Sensitive features, and are therefore likely to be classified accurately.

**Figure S4.**
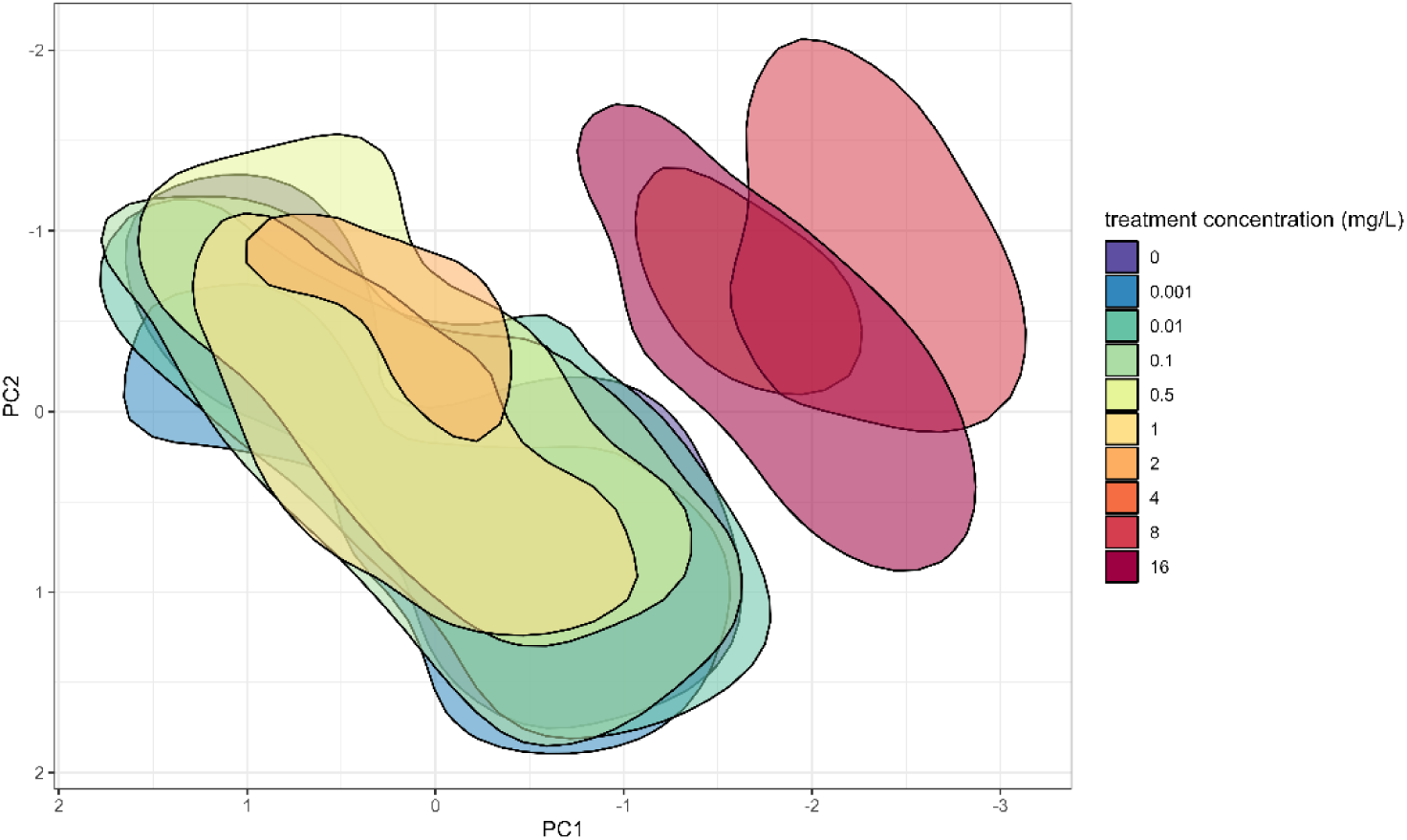
Resistant *E. coli* treated with high concentrations of ciprofloxacin cluster together. This ciprofloxacin-resistant clinical isolate (EC3; MIC = 0.5 mg/L) was treated at varying concentrations of ciprofloxacin for 30 minutes. This principal component analysis shows that features of cells treated at extremely high concentrations of ciprofloxacin (8-, 16-, and 32-times MIC; 4, 8, and 16 mg/L) form a separate cluster from those treated at lower concentrations, even when those concentrations are above the MIC. Geometric shapes are plotted with different colours to show the regions with point density above 0.07.

